# The Impact of Cognitive Load and Encoding Strategies on Prospective Memory in Children with ADHD: Performance and Processing Differences

**DOI:** 10.64898/2026.05.12.26353075

**Authors:** Jiajia Huang, Zhang Lin, Xuanru Wu, Zhaixiang Ye, Yiling Dong, Yixiao Pan

## Abstract

**Introduction:** Prospective memory (PM) deficits in children with attention-deficit/hyperactivity disorder (ADHD) significantly impact academic and daily functioning. Through two experiments, this study investigated how cognitive load and encoding strategies modulate PM performance.

**Methods:** Experiment 1 included 43 children (21 ADHD, 22 typically developing) who completed an n-back task under high and low cognitive load. Experiment 2 included 44 children with ADHD who were randomly assigned to either a standard encoding group or an implementation intention encoding group, also completing the n-back task under both load conditions.

**Results:** Experiment 1 showed that children with ADHD had significantly lower PM accuracy than typically developing peers. Signal detection analysis revealed that this deficit stemmed from a more conservative response bias rather than impaired perceptual sensitivity. Unexpectedly, PM accuracy and perceptual sensitivity were higher under high cognitive load than low load for both groups. Experiment 2 demonstrated that implementation intention encoding significantly enhanced PM accuracy and perceptual sensitivity in children with ADHD, with stable effects across load conditions and no interference with ongoing task performance.

**Discussion:** These findings indicate that PM deficits in children with ADHD reflect a conservative response strategy rather than an inability to detect target cues. Implementation intention encoding provides an effective, load-independent cognitive strategy for enhancing PM performance. These results offer novel insights into the cognitive mechanisms underlying PM deficits in ADHD and provide evidence-based guidance for targeted interventions.

## 1. INTRODUCTION

Prospective memory (PM) refers to the ability to remember and execute intended actions at an appropriate future moment (Einstein & McDaniel, 1990). PM is commonly categorized into two main types: time-based PM, which involves remembering to perform an action at a specific time or after a time interval, and event-based PM (EBPM), which involves remembering to perform an action when a particular external event occurs (Einstein & McDaniel, 1990). A third type, activity-based PM, refers to remembering to perform an action after completing another activity, though this is less frequently studied in experimental paradigms. The present study focuses specifically on event-based PM, as this form of PM closely resembles everyday memory demands—individuals often need to execute intended actions when encountering specific environmental cues (e.g., remembering to buy exam supplies when passing a stationery store). This ecological validity makes EBPM particularly suitable for experimental investigation while maintaining relevance to real-world functioning. Successful PM relies on two components: (1) The prospective component involves noticing the right time or situation to start an intention; (2) The retrospective component involves remembering and carrying out the planned action(Gilles O. Einstein & McDaniel, 1990). Factors influencing PM include age, cognitive resource allocation, and task complexity (Kliegel et al., 2016; Yang et al., 2019). Critically, ongoing tasks (OTs) often interfere with PM because they compete for limited cognitive resources (Matos, Pereira, Albuquerque, & Santos, 2020; Moore, Lampinen, Adams, Nesmith, & Burch, 2022; Ng, Bucks, Loft, Woods, & Weinborn, 2018). For example, when doing multiple tasks at once, PM performance typically declines due to this competition (Chen, Zhang, & Lin, 2022).

A growing body of research has examined prospective memory in children with ADHD. A systematic review by Talbot and colleagues (2017) synthesized six studies and found that while time-based PM deficits were consistently reported, event-based PM deficits were not consistently observed. The review highlighted that the mechanisms underlying these deficits remain unknown, and that no standardized clinical tools or evidence-based interventions for PM in ADHD are currently available.Subsequent studies have extended these findings. Yang et al. (2019) demonstrated that stimulant medication partially ameliorates PM deficits and that cue focality moderates impairment magnitude. Costanzo et al. (2021) further showed that children with ADHD exhibit event-based PM deficits under high working memory load, and that reward improves performance, suggesting a role for motivational factors.More recent research has explored broader implications. Occhionero et al. (2023) found activity-based PM deficits in both children and adults with ADHD using a real-world actigraphy task. Altgassen et al. (2024) further linked everyday PM failures to procrastination, showing that PM mediates the relationship between ADHD symptoms and procrastination behavior. Collectively, this literature establishes PM impairment as a robust feature of ADHD across ages and task types. However, the precise cognitive mechanisms underlying these deficits—particularly the relative contributions of perceptual discrimination versus decision-making processes—remain underspecified. The present study addresses this gap by applying Signal Detection Theory to disentangle these components.

According to diagnostic standards (APA, 2022; WHO, 2010), ADHD is a neurodevelopmental disorder characterized by inattention, impulsivity, and hyperactivity—symptoms that significantly impact daily functioning (Crawford, Kaplan, & Dewey, 2006). Core deficits in ADHD are closely linked to executive function impairments, particularly in working memory, inhibitory control, and task switching (Barkley, 1997; Castellanos & Aoki, 2016; Wang & Feng, 2023). Such deficits may directly contribute to PM difficulties. Talbot and colleagues’ systematic review revealed significant correlations between prospective memory deficits and executive dysfunction in children with ADHD (Talbot, Müller, & Kerns, 2017). These associations are neurobiologically grounded, as executive functions and PM share reliance on prefrontal-striatal circuits (Castellanos & Proal, 2012). Neuroimaging evidence further shows structural and functional abnormalities in brain regions critical for PM (e.g., prefrontal cortex, parietal lobe; Bonath et al., 2018; Castellanos & Aoki, 2016; Nejati & Ghayerin, 2024), which are known to support PM monitoring and retrieval processes (Dianming et al., 2012; Steimke et al., 2013). Beyond the well-documented impact of executive dysfunction, children with ADHD face additional challenges in prospective memory (PM) performance due to core neurocognitive impairments affecting both PM components. According to the Preparatory Attention and Memory (PAM) framework (Smith & Bayen, 2005), the prospective component requires sustained attentional monitoring of environmental cues (Cheie, MacLeod, Miclea, & Visu-Petra, 2017), a process compromised by ADHD-related attention deficits (Wang& Feng, 2023), deficits that reduce monitoring efficiency and intention initiation accuracy (Costanzo et al., 2021). Concurrently, the retrospective component—dependent on successful intention retrieval and execution—is disrupted by working memory impairments characteristic of ADHD, leading to frequent PM failures (Hurks & Hendriksen, 2011). These combined deficits manifest in significant functional impairments including academic underachievement, social interaction difficulties, and reduced emotional self-regulation (Talbot et al., 2017). Consequently, developing targeted interventions to ameliorate PM deficits represents an important research direction with potential clinical applications for this population.

Building on this framework, attentional resources are continuously engaged in environmental monitoring to detect PM cues, consuming sustained working memory resources (Smith & Bayen, 2005). For healthy adults, increased working memory load does not significantly alter PM performance, though it modulates activation in PM-related brain regions (Steimke et al., 2013). In contrast, while high cognitive load typically compromises PM performance in children with ADHD (Cheie et al., 2017), the Multiprocess Theory proposes that salient cues can trigger spontaneous retrieval, whereas non-salient cues require strategic monitoring (Einstein & McDaniel, 2005). Thus, the impact of cognitive load on PM is fundamentally moderated by cue characteristics (Einstein & McDaniel, 2010; Matos et al., 2020). However, empirical evidence examining this contingency in children with ADHD remains limited, warranting systematic investigation.

As previously established, children with ADHD exhibit poorer prospective memory (PM) performance compared to typically developing (TD) peers. This raises the question: Can modified encoding strategies enhance their PM performance? Implementation intention encoding—an effective strategy involving goal specification through “if-then” planning—has demonstrated efficacy in improving PM performance (Rummel, Einstein, & Rampey, 2012). Research indicates this approach significantly enhances PM, particularly the prospective component, by strengthening goal-behavior linkages and reducing reliance on cognitive resources (McDaniel, Howard, & Butler, 2008; Smith, McConnell Rogers, McVay, Lopez, & Loft, 2014). Given that both TD children and those with ADHD have underdeveloped cognitive functions, implementation intentions may reduce cognitive control demands, potentially improving PM task performance.Chen et al. (2022) confirmed that implementation intention encoding improves PM in children with academic difficulties (Chen et al., 2022), while Burkard et al. (2014) observed PM enhancements in cognitively impaired older adults (Burkard, Rochat, Juillerat Van der Linden, Gold, & Van der Linden, 2014), though effects were moderated by working memory capacity. Crucially, these studies did not examine ADHD populations. Specifically, the ‘if-then’ structure of implementation intentions automates the link between PM cues and intended responses by forming a conditional reflex-like association (McDaniel et al., 2008). This automation reduces reliance on depleted executive functions (e.g., sustained monitoring, effortful retrieval) in children with ADHD, thereby compensating for their core cognitive resource deficits and enhancing PM performance. Given that children with ADHD exhibit impaired cognitive resource allocation (Dörrenbächer & Kray, 2019), theoretical convergence suggests that implementation intentions are particularly suited to address these deficits. By automating goal-directed behavior, the strategy can potentially bypass the very executive processes (e.g., sustained monitoring, effortful recall) that are impaired in ADHD, compensating for their core cognitive weaknesses. Furthermore, traditional accuracy metrics cannot dissociate perceptual sensitivity from decision strategies. Signal Detection Theory (SDT) addresses this limitation by separating discriminability (d’) from response bias (c), offering refined analysis of cognitive deficits in ADHD. The “if-then” structure of implementation intentions may simulate SDT’s signal enhancement mechanism by binding goals to actions, potentially increasing target salience and attentional prioritization. However, empirical evidence regarding implementation intention effects on PM in ADHD children remains notably scarce.

In summary, this study proposes the following hypotheses: (1) Children with ADHD will exhibit poorer prospective memory performance compared to typically developing children. Beyond this fundamental impairment, we further hypothesize that (2) both groups will demonstrate better prospective memory under low versus high cognitive load conditions; (3) children with ADHD will display a characteristic signal detection profile featuring comparable perceptual sensitivity (d’) but a more conservative response criterion (c) than TD children (Huang-Pollock et al., 2012; Epstein et al., 2003); and (4) implementation intention encoding will enhance prospective memory in children with ADHD across cognitive load conditions by improving target discriminability without compromising ongoing task performance.

## 2. Experiment 1: Effects of Cognitive Load on PM in ADHD and TD Children

### 2.1. Participants

An a priori power analysis using G*Power 3.1.9.2 indicated a minimum sample of 34 participants to detect a medium effect (f = 0.25, α = .05, power = .80; Cohen, 1988). Given the exploratory nature of this study, we targeted this sample size to detect theoretically and clinically meaningful effects.

To account for potential data loss or attrition, we recruited 43 children in total, which exceeded the minimum required sample size of 34. We recruited 21 children with ADHD (M_age_ = 9.38 years, SD = 1.12) from a hospital outpatient clinic, and 22 typically developing (TD) children (M_age_ = 8.77 years, SD = 2.02) from local elementary schools. All children with ADHD were first-time diagnosed, medication-naïve, and had no prior behavioral interventions. Inclusion criteria for the ADHD group were: (a) first-time clinical diagnosis confirmed by a physician; (b) no prior pharmacological treatment or behavioral interventions; (c) diagnosis verified through standardized assessments (Raven’s Progressive Matrices and SNAP-IV). TD children had no diagnosed psychiatric or neurological conditions per parent report and met the same exclusion criteria regarding sensory impairments and recent experimental participation.

The Raven’s Standard Progressive Matrices (SPM) was used to assess non-verbal reasoning. An independent-samples t-test confirmed no significant group difference in SPM IQ, t(41) = -1.572, p = .124, indicating comparable non-verbal intelligence. ADHD diagnosis was confirmed using the Swanson, Nolan, and Pelham Rating Scale (SNAP-IV), a standardized DSM-based measure. As expected, the ADHD group scored significantly higher than the TD group on both the Inattention subscale, t(41) = 9.167, p < .001, and the Hyperactivity-Impulsivity subscale, t(41) = 11.263, p < .001, validating group classification (see Table 1).

**Table 1.** Demographic and Clinical Characteristics: ADHD vs. TD.

| Characteristic | ADHD1 Group (n = 21) | TD Group (n = 22) | Statistic | p-value |
| --- | --- | --- | --- | --- |
| Demographic |  |  |  |  |
| Age, years (M ± SD) | 9.38 ± 1.12 | 8.77 ± 2.20 | t(41) = -1.572 | 0.124 |
| Gender, male/female (n) | 13/8 | 15/7 | $\chi^2(1) = 0.186$ | 0.666 |
| Clinical Ratings - SNAP-IV (M ± SD) |  |  |  |  |
| Inattention Subscale (Range: 0-3) | 1.63 ± 0.37 | 0.65 ± 0.33 | t(41) = 9.167 | < .001<br>* |
| Hyperactivity-Impulsivity Subscale<br>(Range: 0-3) | 1.33 ± 0.41 | 0.25 ± 0.23 | t(41) = 11.263 | < .001<br>* |
| Cognitive Assessment - Raven's SPM (M ± SE) |  |  |  |  |
| Full-Scale IQ (M ± SD) | 104.19 ± 8.10 | 108.59 ± 10.08 | t(41) = -1.572 | 0.124 |

ADHD=Attention-Deficit/Hyperactivity Disorder; TD=Typically Developing; M =Mean; SD=Standard Deviation; SNAP-IV = Swanson, Nolan, and Pelham Rating Scale, Version IV; SPM = Standard Progressive Matrices. SNAP-IV scores are presented as mean item scores. Raven's SPM IQ scores were converted using age-based normative tables for Chinese children (mean = 100, SD = 15). \* $p < .001$ .

All participants were tested individually in a quiet room. The study was approved by the Ethics Committee of Wenzhou Seventh People’s Hospital (Approval No. EC-KY-202408). Written informed consent was obtained from guardians, and all children provided verbal assent. Participants were naïve to the experimental procedures and received a small gift upon completion.

### 2.2. Method

#### 2.2.1. Design

This study employed a 2 (Group: children with ADHD vs. typically developing children) × 2 (Ongoing task cognitive load: high vs. low) mixed experimental design. Ongoing task cognitive load was manipulated as a within-subjects factor, while group was a between-subjects factor. To control for order effects, the sequence of the cognitive load levels was counterbalanced across participants. The dependent variables assessed were the accuracy and reaction times (RTs) for both the prospective memory task and the ongoing task.

#### 2.2.2. Materials

The experimental materials consisted of disyllabic words randomly selected from the vocabulary lists of primary school Chinese textbooks (Grades 1-6). Specifically, the ongoing task materials included 30 plant-related words and 30 animal-related words. For the prospective memory task, the materials comprised 10 appliance-related words and 10 stationery-related words.

### 2.3. Tasks

Ongoing Task:Under the high cognitive load condition (2-back task), participants were required to judge whether the currently presented word matched the word presented two items back. Under the low cognitive load condition (1-back task), participants judged whether the currently presented word matched the immediately preceding word. For both conditions, participants pressed the ‘F’ key for matching stimuli and the ‘J’ key for non-matching stimuli.

Prospective Memory Task:Participants were instructed to press the ‘P’ key whenever a target word appeared.

### 2.4. Procedure

The formal experimental instructions were presented as follows: “In this task, you will see a sequence of words. Your primary task is to remember each word and compare it with the one that came immediately before it. If the current word is identical to the previous word (e.g., ‘peach’ → ‘peach’), press the ‘F’ key. If the current word is different from the previous word (e.g., ‘peach’ → ‘grass’), press the ‘J’ key. Additionally, and most importantly, whenever you see a word that represents a stationery item (e.g., ‘pen’, ‘ruler’), you must press the ‘P’ key immediately.” Participants were also informed that the example words would not appear in the formal test.

The experimenter explicitly emphasized that the prospective memory (PM) task and the ongoing (n-back) task were equally important and that participants should perform both tasks attentively.

After the instruction phase, participants were asked to repeat the PM task rules to ensure comprehension; all participants correctly repeated the PM instructions before proceeding to the practice block.

Each experimental block comprised 10 target trials for the prospective memory task and 72 trials for the ongoing task. The 10 prospective memory target trials appeared at random positions within the block. Following the PM instruction, the formal test began immediately without any filler or delay task. This design choice was made to maintain task flow and minimize working memory demands, as the entire experimental session lasted approximately 20 minutes—a duration that could be challenging for children with ADHD to sustain attention if additional delay intervals were included.

The stimulus presentation sequence (illustrated in the figure) was as follows: A fixation cross (”+”) was displayed at the center of the screen for 500 ms, followed by the presentation of a word stimulus for 2500 ms. Participants were required to respond to the word stimulus during this 2500 ms period. Immediately after the participant’s response or after 2500 ms elapsed (whichever occurred first), a blank screen was presented, marking the end of the trial and initiating the next trial cycle (see Figure 2). After completing the entire task, participants were again asked to recall the PM instruction (”What were you supposed to do when you saw a stationery item?”). All participants included in the analysis correctly repeated the instruction, confirming that PM failures during the task were not due to forgetting the intention itself. This paradigm (no filler task, 10 PM targets) is consistent with established developmental event-based PM tasks for children (Cheie et al., 2017; Chen et al., 2022), and was used to ensure sufficient trials for reliable SDT analysis while keeping task duration appropriate for children with ADHD.

**Figure 1.**
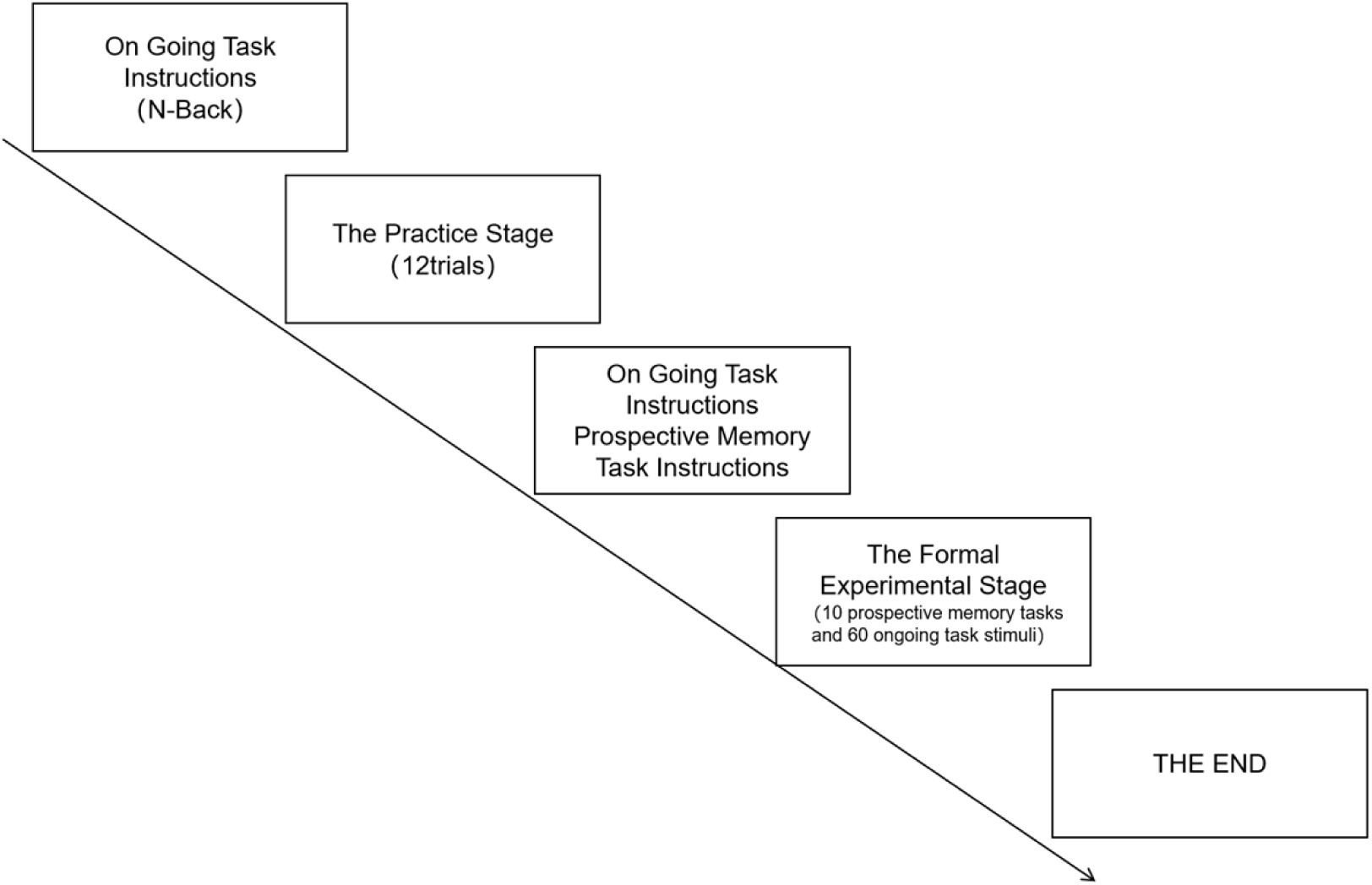
(Experimental Procedure Diagram) Illustrates the standardized study workflow: n-back instruction → 12-trial practice → PM instruction → formal stage (10 PM trials + 60 OT trials) to ensure consistent task execution across participants.

**Figure 2.**
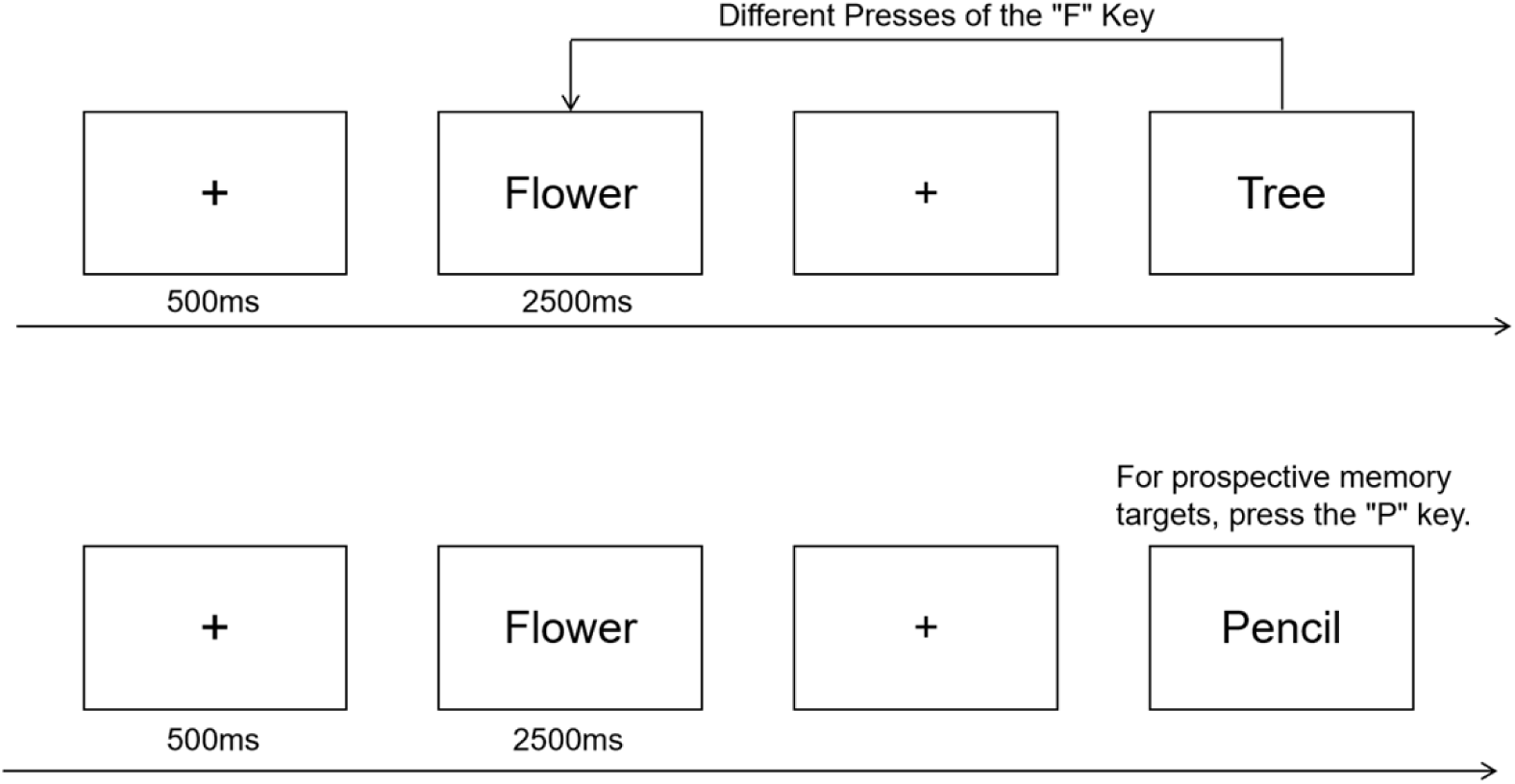
(Stimulus Presentation Flowchart) Depicts the trial sequence: 500-ms fixation cross → 2500-ms word stimulus (for response) → blank screen, clarifying stimulus timing consistency across cognitive load conditions.

### 2.5. Results

Preliminary Data Inspection: preliminary inspection confirmed no floor or ceiling effects (PM: 10–100%; OT: 6–95%). Skewness and kurtosis values for all measures were within acceptable limits (|skewness| < 3, |kurtosis| < 10), indicating approximately normal distributions suitable for parametric analysis.

Data points exceeding ±3 standard deviations were excluded from analysis. All data points fell within ±3 standard deviations of the mean; therefore, no trials were excluded from analysis. The sample comprised 43 participants: 21 children with ADHD and 22 typically developing children. All statistical analyses were performed using SPSS 20.0 (Version 20.0).

Performance on both the prospective memory (PM) and ongoing tasks (OT) was primarily assessed using accuracy rates. The scoring rules were defined as follows:

PM task accuracy = (Number of correct responses to PM target words / Total number of PM target words). Non-responses to PM targets were classified as errors and incorporated into the accuracy analysis, consistent with the requirement for active monitoring in the task

OT accuracy = (Number of correct “same” or “different” judgments / Total number of OT trials)

#### 2.5.1. PM Performance

Prospective memory accuracy and reaction times (RTs,) for typically developing children and children with ADHD under high and low cognitive load conditions are presented in Table 2, Figure 3. A 2 × 2 repeated-measures ANOVA was first conducted on prospective memory accuracy and RTs.

**Figure 3.**
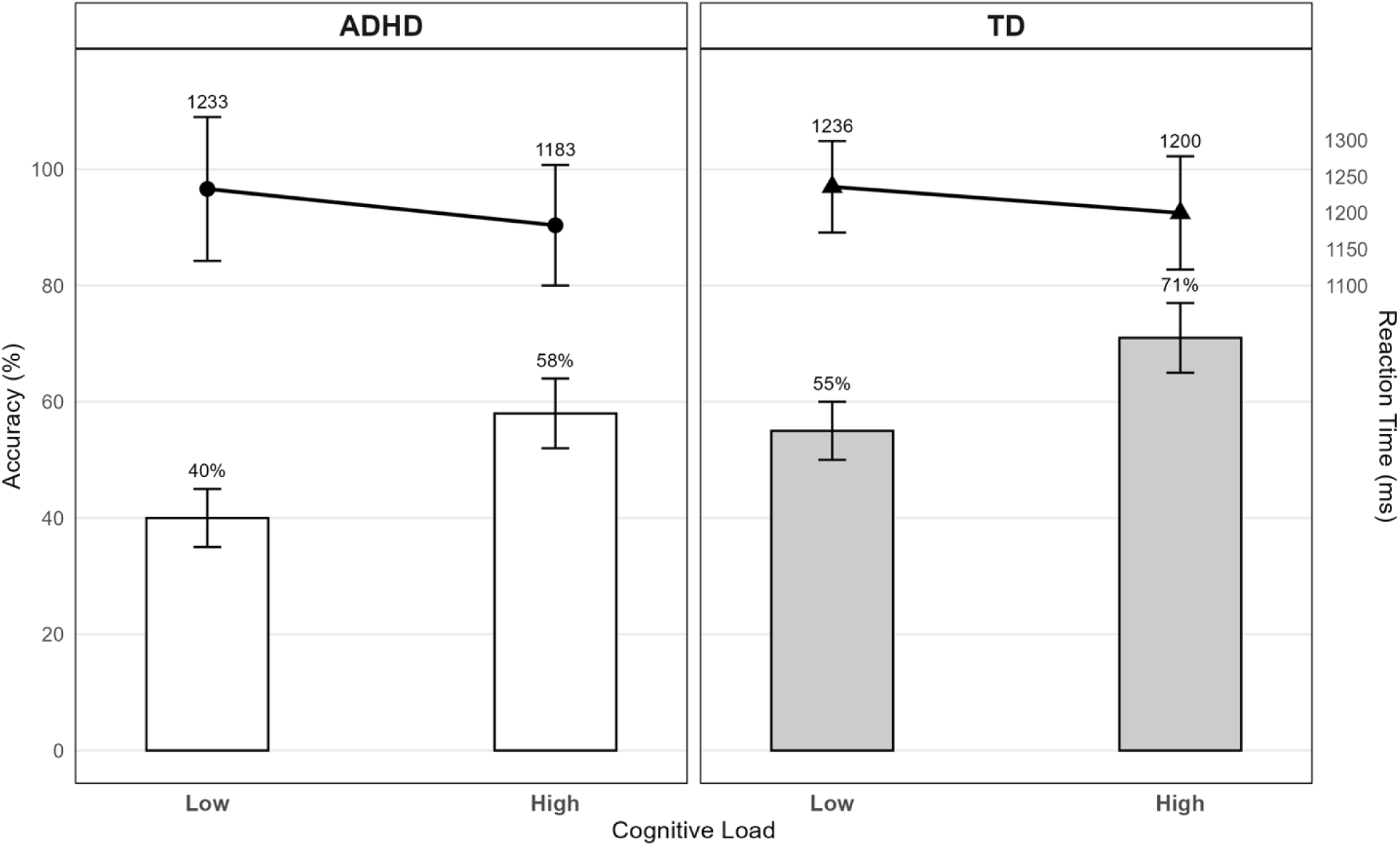
(Prospective Memory Performance: ADHD vs TD Children) Visually compares PM accuracy and RTs: ADHD children have lower PM accuracy than TD peers, and both groups show higher accuracy and faster RTs under high cognitive load.

**Table 2.** Task Performance Across Groups: ADHD vs TD Children.

| cognitive load | Group | PM-ACC(M±SE) | PM-RTs(ms, M±SE) | OT-ACC(M±SE) | OT-RTs(ms, M±SE) |
| --- | --- | --- | --- | --- | --- |
| Low | ADHD | 0.40 ± 0.05 | 1233 ± 99 | 0.62 ± 0.05 | 983 ± 48 |
|  | TD | 0.55 ± 0.05 | 1236 ± 63 | 0.74 ± 0.03 | 1236 ± 63 |
| High | ADHD | 0.58 ± 0.06 | 1183 ± 83 | 0.47 ± 0.06 | 1173 ± 52 |
|  | TD | 0.71 ± 0.06 | 1200 ± 78 | 0.61 ± 0.05 | 1145 ± 61 |

PM = Prospective Memory, OT = Ongoing Task, RTs = Reaction Times. All data are presented as "mean ± standard error". The same notation applies to the following tables.

##### For accuracy

The main effect of cognitive load was significant, *F*(1, 41) = 12.473, *p* = 0 .001, *η²* = 0.233, with both groups showing higher accuracy under high cognitive load than low cognitive load. The main effect of group was significant, *F*(1, 41) = 6.397, *p* = 0.015, *η²* = 0.135, indicating superior accuracy in typically developing children compared to children with ADHD. The cognitive load × group interaction was nonsignificant, *F*(1, 41) = 0.074, *p* = 0.786, *η²* = 0.002.

##### For correct RTs

The main effect of cognitive load was significant, *F*(1, 41) = 8.172, *p* = 0.007, *η²* = 0.166, with faster responses under high cognitive load than low cognitive load. Neither the main effect of group (*F*(1, 41) = 0.697, *p* = 0.409, *η²* = 0.017) nor the cognitive load × group interaction (*F*(1, 41) = 0.197, *p* = 0.659, *η²* = 0.005) reached significance.

#### 2.5.2. OT Performance

To examine the effects of cognitive load levels on ongoing task performance across participant groups (see Table 1, Figure 4), a 2 × 2 repeated-measures ANOVA was similarly conducted for ongoing task accuracy and correct reaction times (RTs).

**Figure 4.**
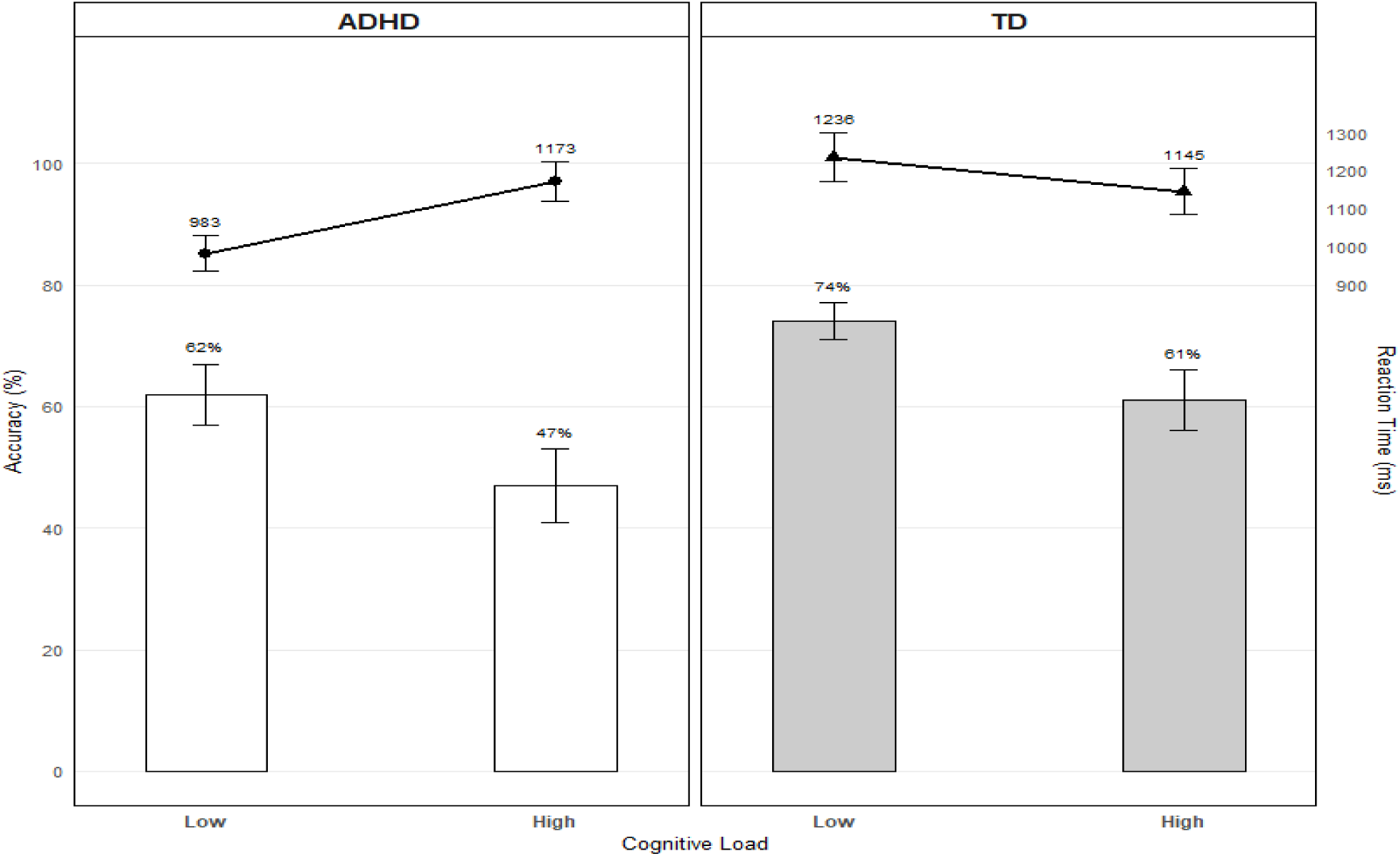
(Ongoing Task Performance: ADHD vs TD Children) Visualizes OT accuracy and RTs: TD children have higher OT accuracy; a load×group interaction is evident (TD faster under low load, ADHD faster under high load).

##### For accuracy

The main effect of cognitive load was significant, *F*(1, 41) = 21.82, *p* < 0.001, *η²* = 0.347, with higher accuracy under low cognitive load than high cognitive load. The main effect of group was significant, *F*(1, 41) = 4.86, *p*= 0.033, *η²* = 0.106, indicating superior accuracy in typically developing children compared to children with ADHD. The cognitive load × group interaction was nonsignificant, *F*(1, 41) = 0.045, *p* = 0.834, *η²* = 0.001.

##### For correct RTs

The main effect of cognitive load was nonsignificant, *F*(1, 41) = 1.82, *p* = 0.185, *η²* = 0.042. The main effect of group approached significance, *F*(1, 41) = 3.74, *p* = 0.060, *η²* = 0.084. Crucially, the cognitive load × group interaction was significant, *F*(1, 41) = 11.276, *p* = 0.002, *η²* = 0.220.

#### 2.5.3. Accuracy-Reaction Time Efficiency Ratio (ART)

To jointly capture both the accuracy and speed of task performance, we computed a standardized ratio of accuracy to reaction time. This metric, referred to as the Accuracy-Reaction Time efficiency ratio (ART), provides an integrated measure of processing efficiency by accounting for the trade-off between speed and accuracy, ART = ACC / RTs (for correct responses).(see Table 3, Figure 5).

**Table 3.** Signal Detection and Efficiency Metrics in ADHD and TD Children Across Task Types load.

| cognitive load | Group | PM-ART | PM-$d'$ | PM-$c$ | OT-ART | OT-$d'$ | OT-$c$ |
| --- | --- | --- | --- | --- | --- | --- | --- |
| Low | ADHD | 0.35 ± 0.06 | 1.85 ± 0.20 | -1.25 ± 0.08 | 0.66 ± 0.08 | 1.93 ± 0.23 | -0.44 ± 0.12 |
|  | TD | 0.47 ± 0.05 | 1.72 ± 0.18 | -0.71 ± 0.05 | 0.61 ± 0.04 | 2.14 ± 0.17 | -0.39 ± 0.11 |
| High | ADHD | 0.53 ± 0.06 | 2.35 ± 0.25 | -0.92 ± 0.08 | 0.42 ± 0.06 | 1.29 ± 0.18 | -0.68 ± 0.18 |
|  | TD | 0.62 ± 0.06 | 2.52 ± 0.25 | -0.65 ± 0.08 | 0.56 ± 0.06 | 1.36 ± 0.17 | -0.69 ± 0.15 |

ART = Accuracy-Reaction Time Efficiency Ratio

**Figure 5.**
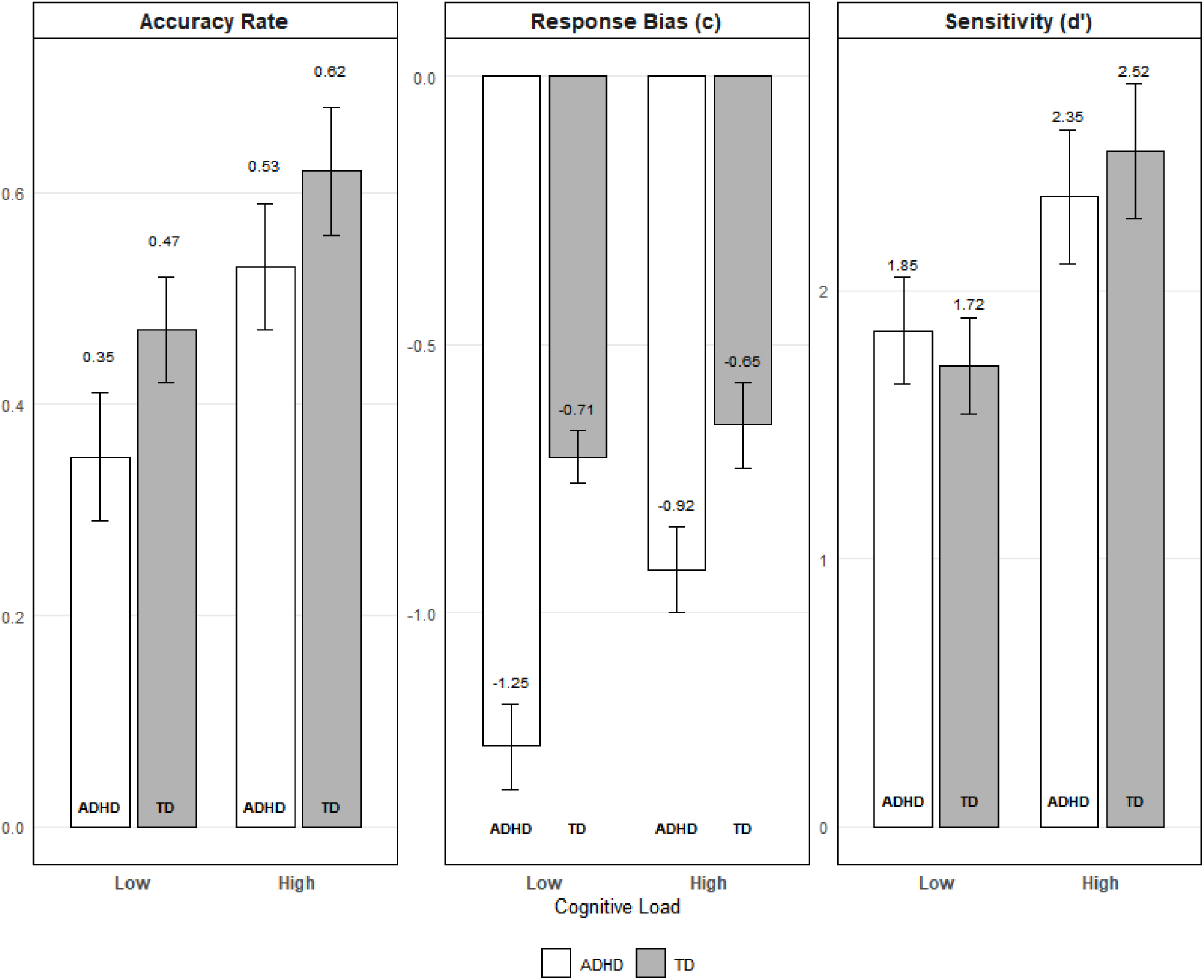
(Processing Efficiency Across Task Types: ADHD vs TD) Shows group differences in ART: both groups have higher PM-ART under high load and higher OT-ART under low load, with no significant between-group differences in efficiency.

##### For PM ART

The main effect of cognitive load was significant, *F*(1, 41) = 15.281, *p* < 0.001, *η²* = 0.272, again showing lower efficiency scores under low cognitive load. The main effect of group was nonsignificant, *F*(1, 41) = 2.473, *p* = 0.124, *η²* = 0.057. The cognitive load × group interaction was nonsignificant, *F*(1, 41) = 0.176, *p*= 0.677, *η²* = 0.004.

##### For OT ART

The main effect of cognitive load was significant, F(1, 41) = 18.995, *p* < 0.001, *η²* = 0.317, with lower efficiency scores under low cognitive load than high cognitive load. The main effect of group was nonsignificant, *F*(1, 41) = 0.36, *p* = 0.552, *η²* = 0.009. Importantly, the cognitive load × group interaction reached significance, *F*(1, 41) = 7.91, *p* = 0.008, *η²* = 0.160.

#### 2.5.4. Signal Detection Analysis

To better assess the influence of decision-making processes, we computed perceptual sensitivity (d’) and response bias (c) indices based on Signal Detection Theory, where d’ = *Z*(Hit rate) – *Z*(False Alarm rate) and c = –0.5 × [*Z*(Hit rate) + *Z*(False Alarm rate)]. The criterion c was interpreted as follows: c = 0 indicates a neutral bias, c > 0 indicates a conservative bias and c < 0 indicates a liberal bias (Macmillan & Creelman, 2005), with results presented in Table 3..

##### For PM perceptual sensitivity (d’)

The main effect of cognitive load was significant, *F*(1, 41) = 12.858, *p* = 0.001, *η²* = 0.239, with higher perceptual sensitivity under high cognitive load than low cognitive load. The main effect of group was nonsignificant, *F*(1, 41) = 0.025, *p* = 0.876, *η²* = 0.001, indicating no significant difference in average perceptual sensitivity (d’) between the ADHD group and the TD group. The cognitive load × group interaction was nonsignificant, *F*(1, 41) = 0.845, *p* = 0.363, *η²* = 0.020..

##### For PM response bias (c)

The main effect of cognitive load was significant, *F*(1, 41) = 7.916, *p* = 0.007, *η²*= 0.162, with a more liberal response bias (lower c-value) under high cognitive load than low cognitive load. The main effect of group was significant, *F*(1, 41) = 31.320, *p* <0 .001, *η²* = 0.433, indicating a more conservative response bias (higher c-value) in the ADHD group compared to the TD group. The cognitive load × group interaction approached significance, *F*(1, 41) = 3.366, *p* = 0.074, *η²* = 0.076.

##### For OT perceptual sensitivity (d’)

The main effect of cognitive load on perceptual sensitivity *d’.* was significant, *F*(1,41)=34.287, *p* <0.001, *η²* =0.455, with the d′-value under low cognitive load being significantly higher than that under high cognitive load. However, neither the main effect of participant group, *F*(1,41)=0.354, *p* = 0.555, *η²* =0.009, nor the interaction between cognitive load and participant group, *F*(1,41)=0.274, *p* =0.604, *η²* = 0.007, was significant for perceptual sensitivity *d’.*.

##### For OT response bias (c)

The main effect of cognitive load was significant, *F*(1,41) =5.899, *p* = 0.020, *η²* =0.126, with the c-value under high cognitive load being significantly lower than that under low cognitive load. The main effect of participant group was not significant, F(1,41)=0.036, *p*=0.850, *η²*=0.001, and the interaction between cognitive load and participant group also failed to reach significance, *F*(1,41)=0.151, *p* =0.700, *η²*=0.004.

### 2.6. Brief Summary

#### Prospective Memory Task Results

Typically developing (TD) children demonstrated significantly higher PM accuracy than children with ADHD. Analysis of Signal Detection Theory indices revealed no significant group difference in perceptual sensitivity (*d’.*). However, children with ADHD exhibited a significantly more conservative response bias (c) than TD children. For both groups, PM accuracy and perceptual sensitivity were significantly higher under high cognitive load compared to low cognitive load, and response times were significantly faster. Both groups also demonstrated a significantly more liberal response bias under high load.

#### Ongoing Task Results

TD children demonstrated significantly higher ongoing task accuracy than children with ADHD. Both groups showed significantly higher accuracy and processing efficiency under low cognitive load compared to high cognitive load. A significant cognitive load × group interaction was found for correct reaction times, with TD children responding faster under low load and ADHD children showing the opposite trend.

The results demonstrate that: (1) Children with ADHD exhibit impaired PM accuracy relative to TD peers, associated with a more conservative response bias but intact perceptual sensitivity; (2) High cognitive load enhances PM performance across groups; (3) Children with ADHD show impaired ongoing task performance and utilize different processing speed strategies under cognitive load.

## 3. Experiment 2: Effects of Encoding and Cognitive Load on PM in ADHD Children

### 3.1. Participants

An a priori power analysis conducted with G*Power 3.1.9.2 indicated a minimum total sample size of 34 was required to detect a medium effect size (*f* = 0.25) at α = 0.05 with 80% power. Two new groups of children with ADHD were recruited, meeting the same inclusion criteria as the ADHD group in Experiment 1. None of these participants had been involved in any previous related experiments. The newly recruited participants were randomly assigned to either the standard encoding group (SE) or the implementation intention encoding group (II) using a random number table method._No outliers exceeding ±3 standard deviations were identified; all 44 participants were included in the final analysis: 21 in the standard encoding group (SE group, *M*_age_= 9.45 years, SD = 1.03); and 23 in the implementation intention encoding group (II group, *M*_age_=9.74 years, SD = 1.25). Baseline comparisons revealed no significant differences between the standard and implementation intention encoding groups in non-verbal intelligence (SPM IQ; *t*(42) =0.115, *p* = 0.537) or ADHD symptoms (SNAP-IV Inattention: *t*(42) =0.285, *p* = 0.552; Hyperactivity-Impulsivity: *t*(42) = -1.263, *p* = 0.213) (see Table 4). The study procedure was approved by the Ethics Committee of Wenzhou Seventh

**Table 4.** Demographic and Clinical Characteristics:Standard vs. Implementation Intention Encoding.

| Characteristic | SE Group (n = 21) | II Group (n = 22) | Statistic | P-value |
| --- | --- | --- | --- | --- |
| Demographic |  |  |  |  |
| Age, years (M ± SD) | 9.45 ± 1.03 | 9.74 ± 1.25 | t(42) = -0.894 | 0.176 |
| Gender, male/female (n) | 14 / 7 | 20 / 3 | $\chi^2(1) = 2.573$ | 0.109 |
| Clinical Ratings - SNAP-IV (M ± SD) |  |  |  |  |
| Inattention Subscale (Range: 0-3) | 1.73 ± 0.33 | 1.71 ± 0.29 | t(42) = 0.285 | 0.552 |
| Hyperactivity-Impulsivity Subscale (Range: 0-3) | 1.21 ± 0.38 | 1.34 ± 0.34 | t(42) = -1.263 | 0.213 |
| Cognitive Assessment - Raven's SPM (M ± SE) |  |  |  |  |
| Full-Scale IQ (M ± Sd) | 106.10 ± 10.78 | 105.74 ± 9.67 | t(42) = 0.115 | 0.537 |

People’s Hospital (EC-KY-202408), and guardian consent was obtained.. All implementation intention group participants were naïve to similar experiments and were tested individually.

Data analysis was performed using SPSS 20.0. After excluding data points exceeding ±3 standard deviations, the final valid sample consisted of 44 participants.

### 3.2. Method

#### 3.2.1. Materials

The same as the materials used in Experiment 1

#### 3.2.2. Design

This study employed a 2 (Group: standard encoding group vs. implementation intention encoding group) × 2 (Ongoing task cognitive load: high vs. low) mixed experimental design. Encoding methods (standard encoding, implementation intention encoding) manipulated as a between-subjects factor, cognitive load (high, low) manipulated as a within-subjects factor. The experimental sequence for the cognitive load conditions was counterbalanced across participants.

### 3.3. Procedure

Except for the differing instructions for the prospective memory task, all other procedures were identical to those in Experiment 1. Implementation Intention Encoding Condition: Participants in this condition received an instruction formulated as an “if-then” plan: “If a stationery word appears, then I press the ‘P’ key immediately!” To enhance the encoding of this plan, participants were instructed to silently repeat the “if-then” statement to themselves while visualizing themselves performing the action. They were then asked to verbally repeated it to ensure comprehension. Standard Encoding Condition: Participants received a standard goal instruction: “When a stationery word appears, press the ‘P’ key immediately!” This instruction specifies the “what” and “when” of the task but does not prompt the formation of a specific “if-then” mental link. This instruction was also verbally repeat by participants during the instruction phase to ensure comprehension, consistent with the procedure in the Implementation Intention Encoding Condition. This paradigm (no filler task, 10 PM targets) is consistent with established developmental event-based PM tasks for children (Cheie et al., 2017; Chen et al., 2022), and was used to ensure sufficient trials for reliable SDT analysis while keeping task duration appropriate for children with ADHD.

### 3.4. Results

Preliminary Data Inspection

Preliminary inspection confirmed no floor or ceiling effects (PM: 10–100%; OT: 12–95%). Skewness and kurtosis values for all measures were within acceptable limits (|skewness| < 3, |kurtosis| < 10), indicating approximately normal distributions suitable for parametric analysis.

#### 3.4.1. PM Performance

For both prospective memory accuracy and reaction time (see Table 5, Figure 6) across encoding conditions and cognitive load levels, separate 2 × 2 repeated-measures ANOVAs were conducted.

**Figure 6.**
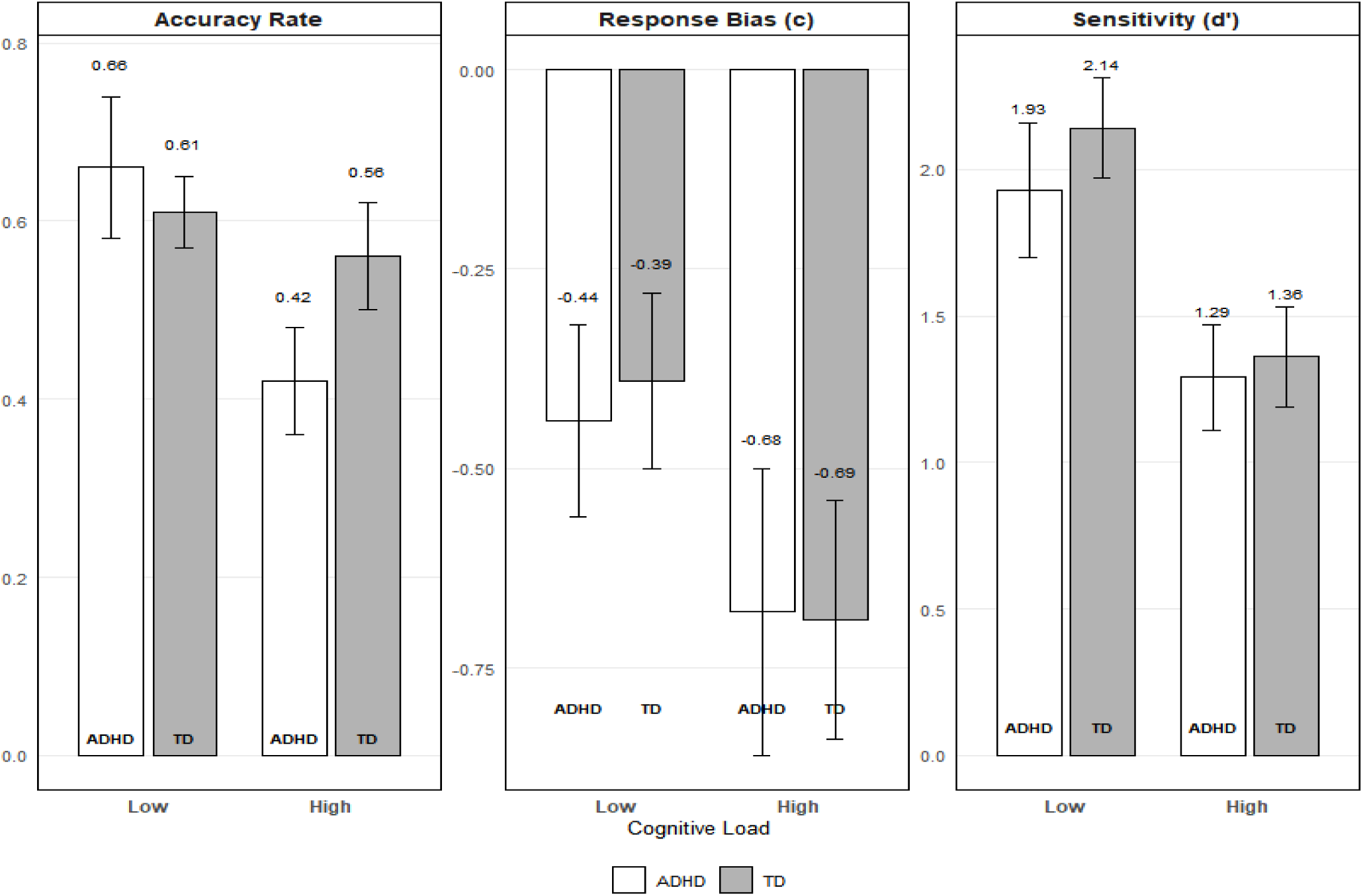
(PM Performance by Encoding Method) Compares PM accuracy and RTs for SE vs. II: II encoding leads to significantly higher PM accuracy and faster RTs in ADHD children, with stable effects across low/high load.

**Table 5.**
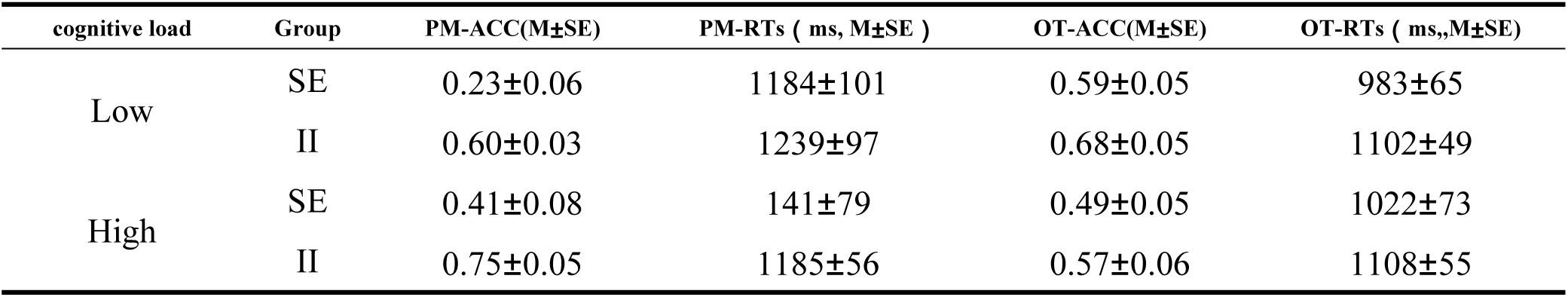
Prospective Memory Performance by Encoding Strategy.

| cognitive load | Group | PM-ACC(M±SE) | PM-RTs ( ms, M±SE ) | OT-ACC(M±SE) | OT-RTs ( ms,,M±SE) |
| --- | --- | --- | --- | --- | --- |
| Low | SE | 0.23±0.06 | 1184±101 | 0.59±0.05 | 983±65 |
|  | II | 0.60±0.03 | 1239±97 | 0.68±0.05 | 1102±49 |
| High | SE | 0.41±0.08 | 141±79 | 0.49±0.05 | 1022±73 |
|  | II | 0.75±0.05 | 1185±56 | 0.57±0.06 | 1108±55 |

##### For accuracy

The main effect of cognitive load was significant, *F*(1, 42) = 19.756, *p* <0 .001, *η²* = 0.320, with higher accuracy under high cognitive load than low cognitive load. The main effect of group was significant, *F*(1, 42) = 24.132, *p* < 0.001, *η²* = 0.365, indicating superior accuracy in the II group compared to the SE group. The cognitive load × group interaction was nonsignificant, *F*(1, 42) = 0.147, *p* = 0.703, *η²* = 0.003,

##### For correct RTs

The main effect of cognitive load was nonsignificant, *F*(1, 42) = 1.445, *p* = 0.236, *η²*= 0.033, with no significant difference in reaction time between low cognitive load and high cognitive load. The main effect of group was significant, *F*(1, 42) = 11.099, *p* = 0.002, *η²*= 0.209, indicating that the average reaction time differed significantly between the SE group and the II encoding group (B). The cognitive load × group interaction approached significance, *F*(1, 42) = 3.922, *p* = 0.054, *η²* = 0.085,

#### 3.4.2. OT Performance

To examine the effects of cognitive load on ongoing task performance in children with ADHD, separate 2 × 2 repeated-measures ANOVAs were conducted for both accuracy and reaction time (see Table 5, Figure 7).

**Figure 7.**
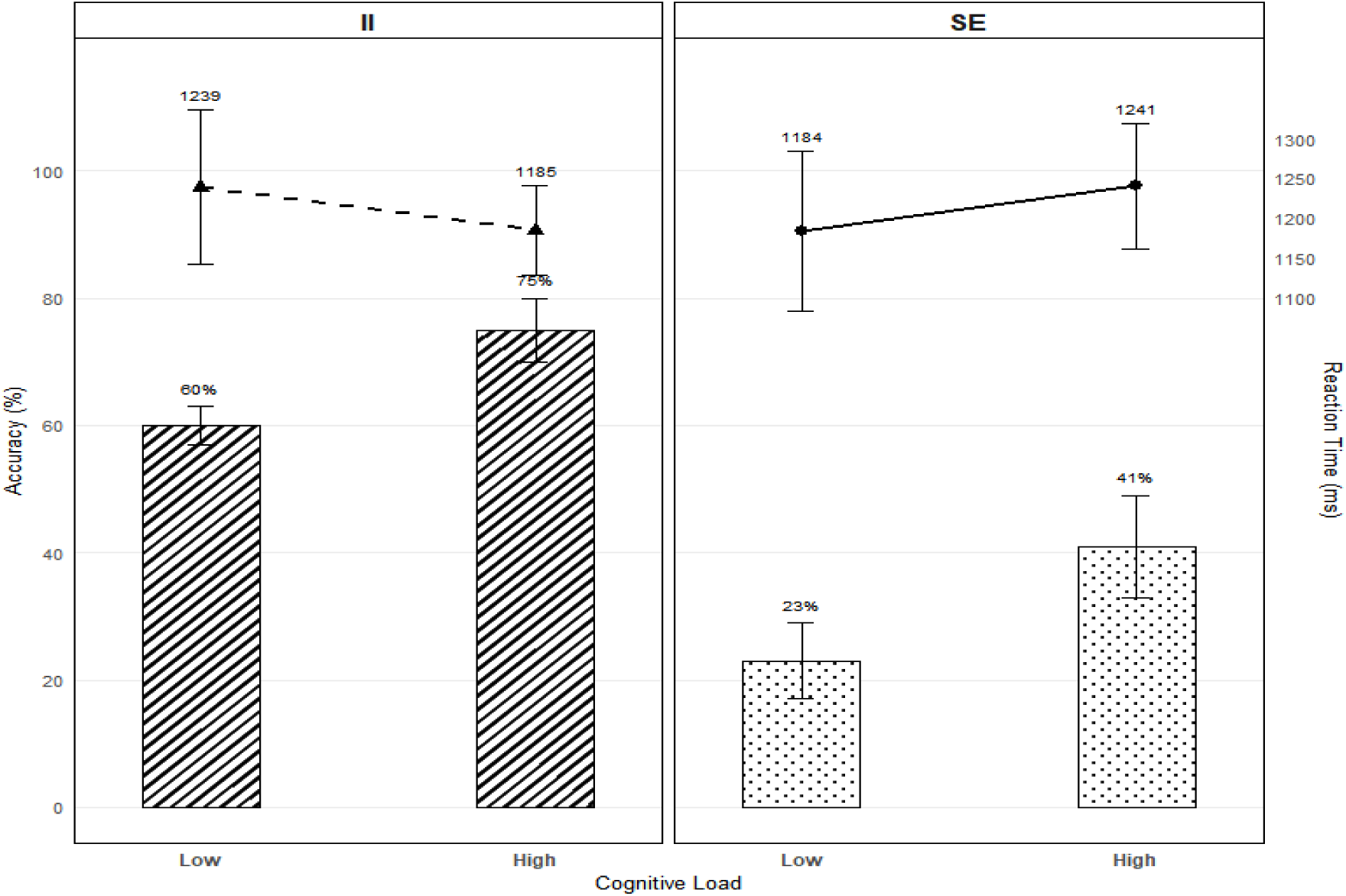
(Prospective Memory Performance by Encoding Method) Visualizes OT accuracy and RTs for SE vs. II: no significant differences in OT performance between encoding groups, confirming II encoding does not interfere with ongoing tasks.

##### For accuracy

The main effect of cognitive load was significant, *F*(1, 42) = 19.315, *p* <0 .001, *η²* = 0.315, with higher accuracy under low cognitive load than high cognitive load. The main effect of group was nonsignificant, *F*(1, 42) = 1.547, *p* = 0.220, *η²* = 0.036, indicating no significant difference in accuracy between the SE group and the II encoding group. The cognitive load × group interaction was nonsignificant, *F*(1, 42) = 0.012, *p* = 0.912, *η²* = 0.000..

##### For correct RTs

The main effect of cognitive load was nonsignificant, *F*(1, 42) = 1.55, *p* = 0.220, *η²* = 0.036,. The main effect of group was also nonsignificant, *F*(1, 42) = 0.625, *p* = 0.434, *η²* = 0.015. The cognitive load × group interaction was nonsignificant, *F*(1, 42) = 0.915, *p* = 0.344, *η²* = 0.021.

#### 3.4.3. Accuracy-Reaction Time Efficiency Ratio (ART)

To better capture the trade-off between accuracy and speed in task performance, we also computed ART,see the Table6, Figure 8 below for detail.

**Figure 8.**
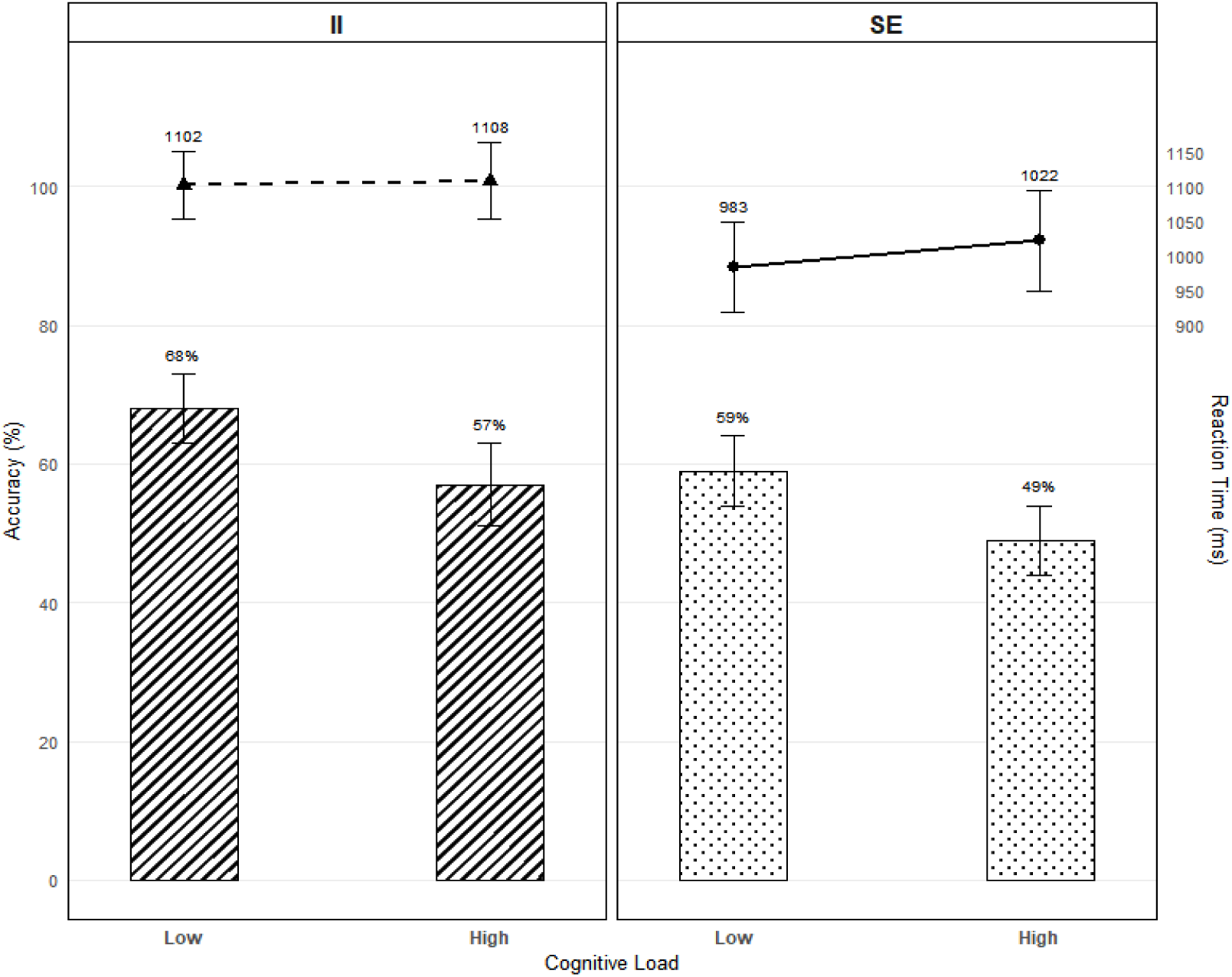
(Processing Efficiency (ART) by Encoding Strategy Across Tasks) Shows ART differences: II group has higher PM-ART than SE group in ADHD children, with no group differences in OT-ART, demonstrating selective PM efficiency improvement via II encoding.

**Figure 9.**
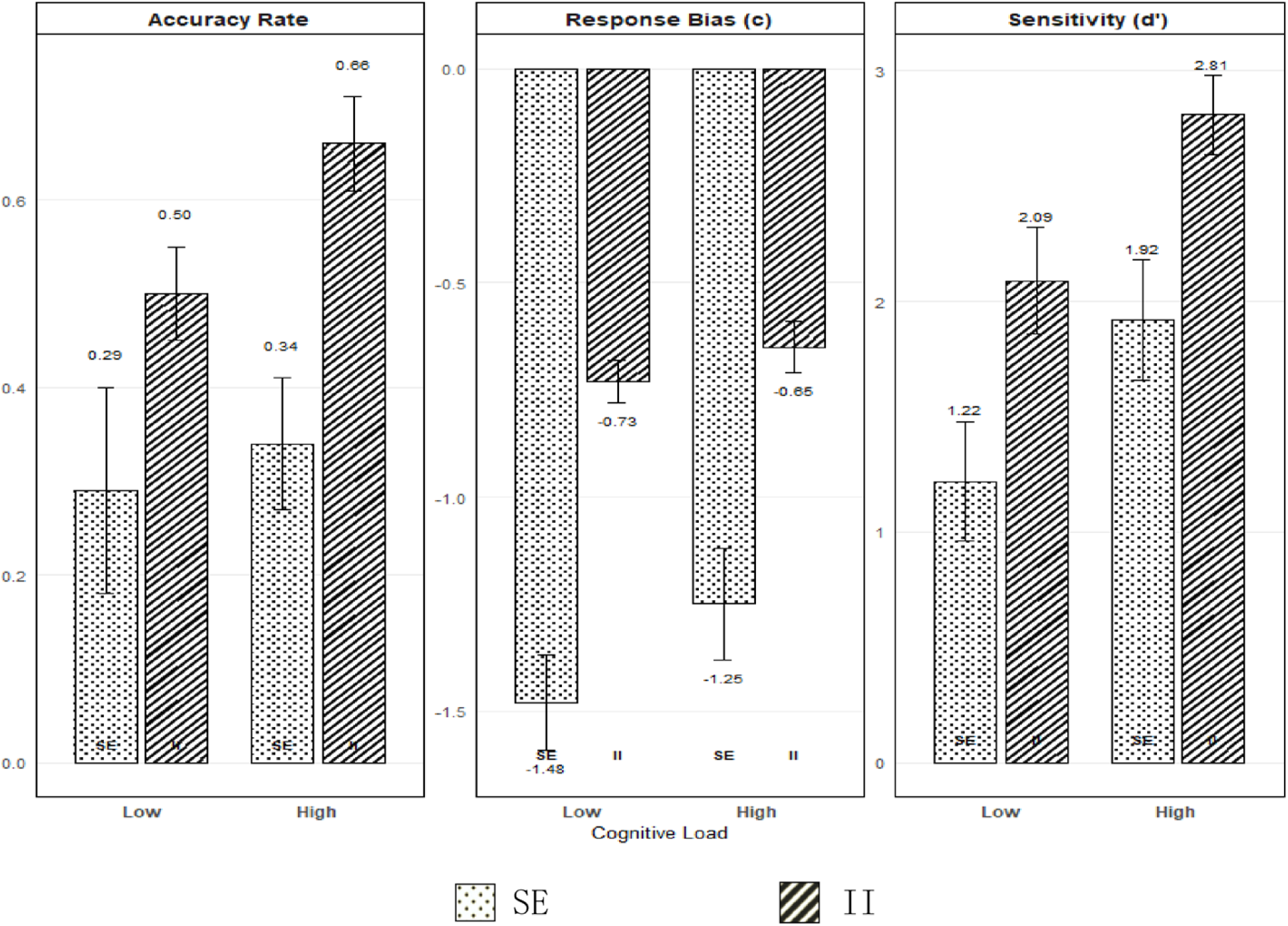
Prospective Memory: Signal Detection and Efficiency Metrics in SE and II Group Across Task Types.

**Figure 10.**
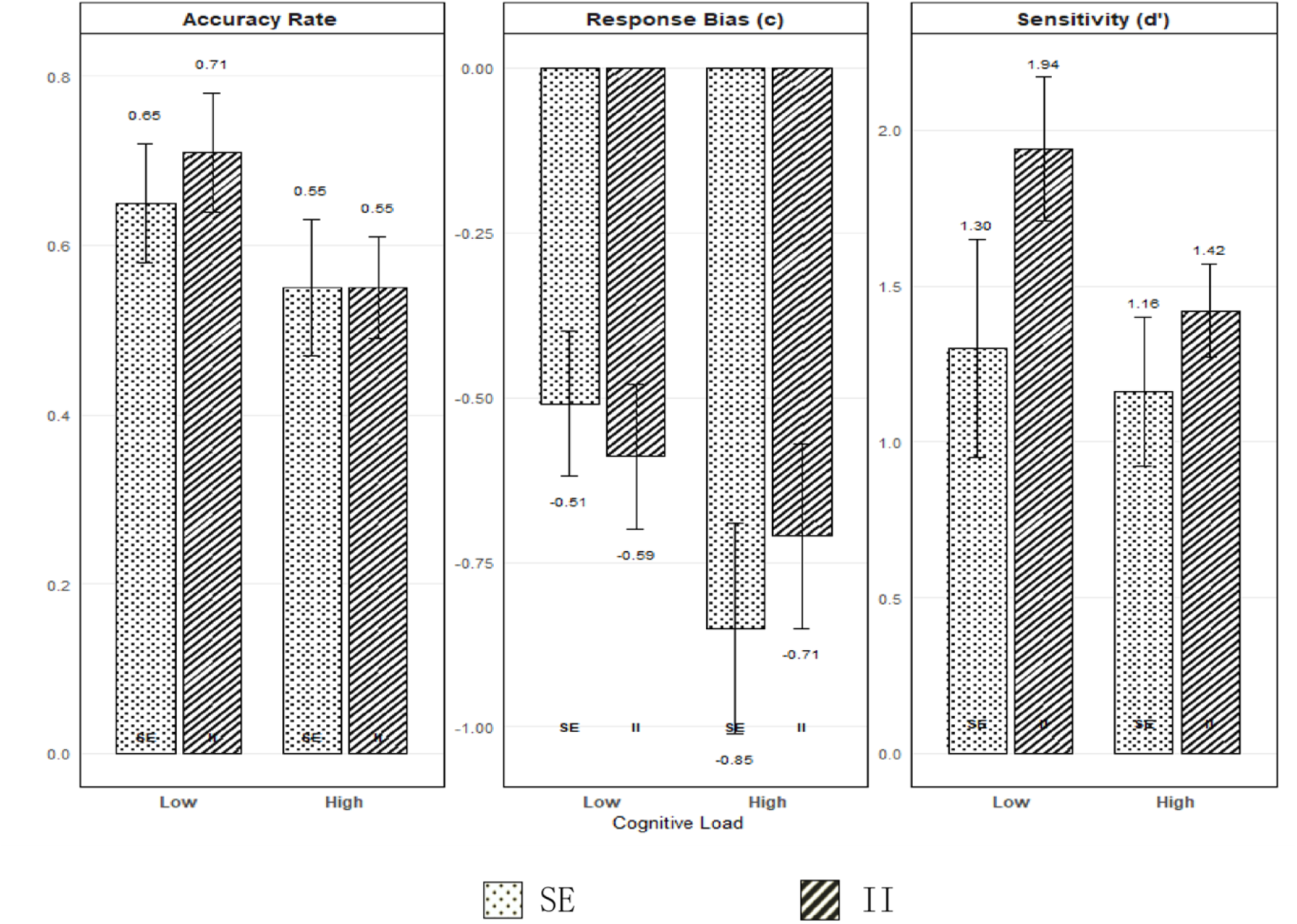
Ongoing Task: Signal Detection and Efficiency Metrics in SE and II Group Across Task Types.

##### For PM ART

The main effect of cognitive load was nonsignificant, *F*(1, 42) = 2.351, *p* = 0.133, *η²* = 0.053. The main effect of group was significant, *F*(1, 42) = 10.834, *p* = 0.002, *η²* = 0.205, indicating superior ART in the II encoding group compared to the SE group. The cognitive load × group interaction was nonsignificant, *F*(1, 42) = 0.636, *p* =0 .430, *η²* = 0.015,

##### For OT ART

The main effect of cognitive load was significant, *F*(1, 42) = 13.156, p =0 .001, *η²* = 0.239, with higher ART under low cognitive load than high cognitive load. The main effect of group was nonsignificant, *F*(1, 42) = 0.107, *p* = 0.745, *η²* = 0.003, indicating no significant difference in average ART between the SE group and the II encoding group. The cognitive load × group interaction was nonsignificant, *F*(1, 42) = 0.484, *p* = 0.491, *η²* = 0.011..

#### 3.4.4. SDT Analysis

To better assess the influence of decision-making processes, we computed perceptual sensitivity (d’) and response bias (c) indices, with results presented in Table 6.

**Table 6.** Cognitive Processing Metrics Across Tasks and Encoding Conditions.

| cognitive load | Group | PM-ART | PM-*d'* | PM-c | OT-ART | OT-*d'* | OT-c |
| --- | --- | --- | --- | --- | --- | --- | --- |
| Low | SE | 0.29±0.11 | 1.22±0.26 | -1.48±0.11 | 0.65±0.07 | 1.30±0.35 | -0.51±0.11 |
|  | II | 0.50±0.05 | 2.09±0.23 | -0.73±0.05 | 0.71±0.07 | 1.94±0.23 | -0.59±0.11 |
| High | SE | 0.34±0.07 | 1.92±0.26 | -1.25±0.13 | 0.55±0.08 | 1.16±0.24 | -0.85±0.16 |
|  | II | 0.66±0.05 | 2.81±0.17 | -0.65±0.06 | 0.55±0.06 | 1.42±0.15 | -0.71±0.14 |

##### For PM perceptual sensitivity (*d’*)

The main effect of cognitive load was significant, *F*(1, 42) = 18.650, *p* <0 .001, *η²* = 0.308, with higher perceptual sensitivity under high cognitive load than low cognitive load. The main effect of group was significant, *F*(1, 42) = 10.546, *p* = 0.002, *η²* = 0.201, indicating superior perceptual sensitivity (*d’*) in the II encoding group (B) compared to the SE group. The cognitive load × group interaction was nonsignificant, *F*(1, 42) = 0.004, *p* = 0.951, *η²* = 0.000.

##### For PM response bias (c)

The main effect of cognitive load was significant, *F*(1, 42) = 9.521, *p* = 0.004, *η²* = 0.185, with a more liberal response bias (lower c-value) under high cognitive load than low cognitive load. The main effect of group was nonsignificant, *F*(1, 42) = 0.019, *p* = 0.892, *η²* = 0.000. The cognitive load × group interaction was nonsignificant, *F*(1, 42) = 1.825, *p* = 0.184, *η²* = 0.042.

##### For OT perceptual sensitivity (d’)

The main effect of cognitive load was significant, *F*(1, 42) = 4.598, *p* = 0.038, *η²* = 0.099, with higher perceptual sensitivity under low cognitive load than high cognitive load. The main effect of group was nonsignificant, *F*(1, 42) = 2.117, *p* = 0.153, *η²* = 0.048. The cognitive load × group interaction was nonsignificant, *F*(1, 42) = 1.603, *p* = 0.212, *η²* = 0.037.

##### For OT response bias (c)

The main effect of cognitive load was significant, *F*(1, 42) = 9.521, *p* = 0.004, *η²* = 0.185, with a more liberal response bias (lower c-value) under high cognitive load than low cognitive load. The main effect of group was nonsignificant, *F*(1, 42) = 0.019, *p* = 0.892, *η²* = 0.000. The cognitive load × group interaction was nonsignificant, *F*(1, 42) = 1.825, *p* = 0.184, *η²* = 0.042.

### 3.5. Brief Summary

#### Prospective Memory Task Results

The implementation intention encoding group demonstrated significantly higher PM accuracy, processing efficiency (ART), perceptual sensitivity (d′), and a more liberal response bias (c) than the standard encoding group. PM accuracy and perceptual sensitivity were significantly higher under high cognitive load compared to low cognitive load for both encoding groups. The response bias was significantly more liberal under high load for both groups. No significant interactions between cognitive load and encoding group were found for any PM measure.

#### Ongoing Task Results

No significant differences in accuracy, processing efficiency (ART), or perceptual sensitivity (d′) were found between the implementation intention and standard encoding groups. Both encoding groups showed significantly higher accuracy and processing efficiency under low cognitive load compared to high cognitive load.

The results demonstrate that: (1) Implementation intention encoding enhances PM performance across multiple metrics in children with ADHD; (2) The beneficial effect of implementation intention encoding is maintained across different cognitive load levels; (3) Implementation intention encoding does not affect ongoing task performance..

## 4. DISCUSSION

This study examined the effects of cognitive load and encoding methods on prospective memory and ongoing task performance in children with ADHD through two experiments. Experiment 1 revealed that children with ADHD showed significantly poorer performance in both prospective memory and ongoing tasks compared to typically developing children, with cognitive load exerting differential effects across task types. Experiment 2 further demonstrated that implementation intention encoding significantly enhanced prospective memory performance in children with ADHD, while exerting no significant effect on ongoing task performance. The discussion will focus on three key aspects: the characteristic task performance patterns in ADHD children, the mechanistic role of cognitive load, and the intervention effectiveness of implementation intention encoding.

### 4.1. Cognitive and Neural Mechanisms of PM Deficits in ADHD

Results from Experiment 1 indicated that children with ADHD exhibited significantly impaired prospective memory (PM) performance compared to their typically developing (TD) peers, thereby supporting Hypothesis 1. These findings align with a growing body of evidence documenting event-based PM difficulties in children with ADHD under cognitively demanding conditions (Yang et al., 2019; Costanzo et al., 2021; Talbot et al., 2017). This finding of impaired event-based PM in a high-demand n-back context aligns with the executive function challenges characteristic of ADHD. The observed deficit is further clarified by the signal detection analysis. While perceptual sensitivity (d’)—the ability to distinguish PM targets from non-targets—was comparable between groups, children with ADHD adopted a significantly more conservative response bias (c). In practical terms, this means that children with ADHD were equally capable of detecting PM cues when they appeared, but were more hesitant to respond, requiring greater certainty before acting. This pattern of intact discriminability (*d’.*) but more conservative response bias (c) in ADHD is consistent with decision-making patterns observed in other cognitive tasks involving uncertainty and resource competition (Huang-Pollock et al., 2012; Epstein et al., 2003). This pattern indicates that their PM impairment primarily stemmed from a decision-making reluctance to respond to potential targets, rather than an inherent inability to perceive them.

Two primary mechanisms, substantiated by our experimental data, may underlie these difficulties. First, executive function impairments characteristic of ADHD likely contribute to these deficits. This interpretation is in line with our ongoing task (OT) results, which revealed significantly poorer working memory performance in children with ADHD compared to TD controls. This direct behavioral evidence aligns with Wang et al.’s (2023) neurocognitive-behavioral developmental pathways linking ADHD symptoms to executive dysfunction (Wang & Feng, 2023), and is particularly relevant given the established relationship between PM and executive functions (Schnitzspahn, Stahl, Zeintl, Kaller, & Kliegel, 2013).

Second, neurodevelopmental abnormalities may play a crucial role. The observed performance pattern—particularly the generalized deficits across both PM and OT tasks—is consistent with models of prefrontal cortex (PFC) dysfunction in ADHD. Evidence indicates delayed or insufficient development of this region (particularly BA10) in children with ADHD (Mack, 2012; Shaw et al., 2013; Wang& Feng,2023), which supports critical functions including attentional control and task switching (Simons & Spiers, 2003). In our paradigm, such neural deficits could help explain why children with ADHD struggled to efficiently allocate cognitive resources for simultaneous PM target detection and OT maintenance, as reflected in their overall lower accuracy profiles.

This dissociation between CPT and PM paradigms—reduced d’ in CPT versus intact d’ but conservative c in PM—suggests that the nature of ADHD-related cognitive deficits depends critically on task characteristics. CPT tasks typically require rapid, frequent responses and tap into sustained attention, whereas our PM task embedded targets within an ongoing cognitive task, placing greater demands on working memory. Under these conditions, children with ADHD may adopt a cautious response strategy to compensate for perceived uncertainty, even when their basic perceptual abilities are intact.

Alternative interpretations should also be considered. The conservative response bias could reflect reduced confidence in memory for the PM intention, leading to hesitation when potential targets appear—an interpretation consistent with the working memory impairments observed in our OT data. Additionally, motivational factors may play a role: children with ADHD might be more sensitive to the perceived costs of errors, particularly in a task where PM and OT are emphasized as equally important. Future studies could test these possibilities by including trial-by-trial confidence ratings, manipulating error costs, or examining whether the bias persists when response criteria are made more explicit.

Our OT data revealed not only lower accuracy in ADHD children across loads, but also a dissociative processing speed pattern: TD children responded faster under low load, whereas ADHD children showed the opposite trend. The significant cognitive load × group interaction for OT correct RTs reflects dissociable resource allocation strategies between the two groups. Typically developing (TD) children exhibited the typical “load-slowing” effect—responding more quickly under low cognitive load and slowing down under high load—which aligns with how cognitive resources are generally consumed by increased task demand. In contrast, children with ADHD showed the opposite pattern: their ongoing task response speed improved under high cognitive load compared to low load. This deviation from the conventional load effect suggests ADHD children prioritize ongoing task (OT) performance when cognitive demand rises, likely as a compensatory effort to maintain OT accuracy—even if this comes at the cost of resources allocated to prospective memory (PM). It is also possible that the low-load (1-back) task was sufficiently easy that some children became bored or less engaged, resulting in slower responses. This divergent pattern aligns with Barkley’s (1997) theory that children with ADHD exhibit inflexible cognitive resource allocation: they struggle to balance dual-task (PM + OT) demands under low load, but shift to focusing on the more structured OT task when load increases, using speed enhancement as a compensatory strategy. This dissociation in resource allocation strategies converges with Freer’s (2011) report of ADHD-TD performance disparities under high cognitive load, and substantiates executive function limitations in ADHD regarding dynamic resource allocation. These findings thereby support Barkley’s (1997) theoretical position that inattention in ADHD fundamentally reflects executive dysfunction. According to optimal stimulation theory (Zentall & Zentall, 1983), children with ADHD are chronically underaroused at typical levels of environmental stimulation and require higher levels of input to maintain optimal arousal. This account can explain the counterintuitive finding of better PM performance under high cognitive load.

### 4.2. Bidirectional Regulatory Effects of Cognitive Load

Experiments 1 and 2 collectively demonstrate that cognitive load significantly influences task performance in both children with ADHD and typically developing children, though its effects manifest differentially across task types. For ongoing tasks, accuracy, ART, and perceptual sensitivity (d’) were consistently superior under low cognitive load conditions—aligning with established findings(Harrison, Mullet, Whiffen, Ousterhout, & Einstein, 2014). This pattern reflects reduced available cognitive resources compromising performance during high cognitive load.

Contrastingly, prospective memory performance revealed higher accuracy and ART values under high cognitive load conditions, contradicting both our hypotheses and prior research(Chen et al., 2022; Harrison et al., 2014). We propose that this paradoxical enhancement can be explained by a strategic reallocation of limited cognitive resources. Under high cognitive load, as the ongoing n-back task becomes exceedingly difficult, participants may have adopted a compensatory strategy of ‘sacrificing OT to protect PM,’ directing their scarce resources toward the prospective memory task. This reallocation resulted in an apparent boost in PM performance, but at the cost of diminished OT accuracy, a pattern that aligns with and clarifies our subsequent findings on the inflexible resource allocation characteristic of ADHD.

Notably, our finding that high cognitive load enhances PM performance contradicts prior studies reporting load-induced PM impairment (e.g., Harrison et al., 2014). This discrepancy can be explained by task characteristics: our PM task was focal (targets belonged to a distinct category, ‘stationery’), whereas prior studies used non-focal PM tasks (targets defined by arbitrary features). According to the Multiprocess Theory (Einstein & McDaniel, 2005), focal PM relies on spontaneous retrieval (less resource-dependent), and high load may prioritize salient focal cues by redirecting limited resources away from the demanding OT (n-back), thereby enhancing PM detection. We confirm no data coding errors (e.g., target word definition, response key assignment were consistent across load conditions), supporting the robustness of this paradoxical effect.

Further support for this interpretation comes from signal detection analyses: high cognitive load significantly improved perceptual sensitivity (d’) and shifted response bias (c) to be more liberal, suggesting that load enhances PM via both improved target discrimination and reduced response hesitation (Castellanos & Proal, 2012). Unlike studies using fixed-sequence prospective targets, our randomized target presentation required continuous task-switching between ongoing and prospective demands. When prospective cues lack salience, working memory resources must actively ‘scan’ the environment (Matos et al., 2020). Under high cognitive load, participants may prioritize the relatively simpler prospective task (Yang et al., 2019) to minimize working memory updating costs associated with frequent task-switching, thereby enhancing prospective performance.

Second, signal detection theory (SDT) analyses provided critical insights into the strategic adaptations underlying the behavioral outcomes. Critically, children with ADHD displayed load-dependent shifts in response strategies that diverged from their typically developing (TD) peers in decision-making bias rather than target discrimination ability. Under low cognitive load, ADHD children exhibited a more conservative response tendency—reluctant to initiate responses to potential PM targets—even though their capacity to distinguish PM targets from non-targets was comparable to TD children. This pattern highlights that ADHD-related PM deficits stem from strategic response adjustments, not impaired perceptual processing of task cues. In stark contrast, high cognitive load triggered a compensatory shift in ADHD children toward a more liberal response bias, accompanied by improved target detection sensitivity that remained aligned with TD children’s performance under the same load condition. This adaptation suggests that the high-demand context redirected limited cognitive resources toward prioritized PM targets, a paradoxical enhancement supported by neurocognitive accounts of compensatory up-regulation under demanding conditions (Sarter, Givens, & Bruno, 2001).

Typically developing children, conversely, maintained stable liberal responding across both load conditions, reflecting robust and consistent monitoring abilities that required no strategic reshaping. A notable trend emerged in the marginal cognitive load × group interaction for response bias, with ADHD children showing a more pronounced shift toward liberal responding when load increased compared to TD children. This suggests that ADHD children’s decision strategies are more sensitive to cognitive demand, potentially representing a context-dependent compensatory mechanism to overcome inherent attentional limitations—one that normalizes their response pattern without incurring excessive false alarm risks. These findings align with SDT frameworks linking ADHD-related cognitive differences to dysregulation within prefrontal-parietal networks (Castellanos & Proal, 2012), but extend prior accounts by emphasizing that the key divergence lies in response bias regulation rather than perceptual sensitivity deficits.

### 4.3. Intervention Effects of Implementation Intention Encoding

Experiment 2 demonstrated that implementation intention encoding significantly enhanced both accuracy and perceptual sensitivity (d’) in prospective memory tasks among children with ADHD. This enhancement in perceptual sensitivity is particularly noteworthy, as it indicates that the ‘if-then’ formulation not only improved overall performance but fundamentally sharpened children’s ability to discriminate PM targets from non-targets. This finding aligns with established research: Chen et al. (2022) observed improved prospective memory performance through implementation intentions in academically underachieving students(Chen et al., 2022), while Smith et al. (2014) empirically validated this enhancement effect across three non-focal prospective memory experiments(Smith et al., 2014). These results collectively support the theoretical proposition that implementation intentions strengthen goal-behavior linkages to facilitate prospective remembering(Rummel et al., 2012).

Mechanistically, the ‘if-then’ formulation appears to help children with ADHD establish clearer target representations while reducing cognitive conflict during intention retrieval (Chen et al., 2022; Rummel et al., 2012). Our SDT results provide direct support for this mechanism, showing that implementation intentions specifically enhanced perceptual sensitivity (d’) without significantly altering response bias (c). Critically, our findings demonstrate that implementation intention encoding provided a significant benefit to prospective memory performance across both low and high cognitive load conditions, as indicated by the strong main effect of encoding. This suggests that II serves as an effective cognitive scaffold for children with ADHD, compensating for executive function deficits by streamlining the intention retrieval process regardless of concurrent load levels. Although the main effect of encoding was significant, we observed a descriptive trend: the improvement in PM performance under low cognitive load was numerically larger than that under high load. This may suggest that when cognitive resources are relatively abundant (low load), children with ADHD can more fully utilize the implementation intention strategy. However, as the load × encoding interaction was not statistically significant, this interpretation requires caution and should be verified in future research. Thus, the beneficial effect of implementation intentions remains stable across varying cognitive load conditions, rather than being more effective in low-load contexts. This interpretation is consistent with our efficiency (ART) data, which showed more substantial gains under low load conditions, suggesting that adequate cognitive resources may be important for maximizing the strategy’s effectiveness.

Moreover, implementation intention encoding showed no significant effects on ongoing task performance, as evidenced by comparable accuracy, reaction times, and efficiency ratios between encoding groups across both load conditions. This selective benefit pattern corroborates prior evidence that implementation intentions primarily optimize goal-directed behaviors rather than fundamental cognitive processes(Rummel et al., 2012). Future research could explore integrating implementation intention encoding with complementary cognitive training approaches to comprehensively enhance multitasking capabilities in children with ADHD.

In summary, the influence of cognitive load on prospective memory in children with ADHD reveals a complex but interpretable pattern. The high cognitive load condition triggered a compensatory adaptation characterized by a liberalized response bias that enhanced PM detection—however, this passive adjustment occurred alongside compromised ongoing task accuracy. In contrast, implementation intention encoding provided an active, top-down cognitive scaffold that significantly improved PM performance across both load conditions, primarily by enhancing perceptual sensitivity. While this strategic intervention remained effective regardless of cognitive load, the descriptively larger improvement under low load suggests that adequate cognitive resources may facilitate optimal strategy implementation. Collectively, these findings underscore that PM deficits in ADHD fundamentally reflect both scarcity and inefficient allocation of cognitive resources. While demanding environments can elicit rudimentary compensatory mechanisms, they may simultaneously constrain the application of refined cognitive strategies. Therefore, effective intervention requires not only providing strategic tools like implementation intentions but also carefully considering the cognitive demands of the execution environment to optimize both resource availability and allocation efficiency.

## 5. Conclusion

This study examines the impact of cognitive load on prospective memory in children with ADHD and assesses whether implementation intention encoding can enhance their prospective memory performance. The findings yielded the following key conclusions:

1. Children with ADHD demonstrated significant impairments in both ongoing task accuracy and prospective memory performance compared to typically developing peers. Signal detection analysis revealed that this deficit was characterized by a more conservative response bias rather than impaired perceptual sensitivity.
2. Contrary to conventional expectations, prospective memory performance was superior under high cognitive load conditions compared to low cognitive load across both participant groups, accompanied by a significant shift toward more liberal response criteria under high load.
3. Implementation intention encoding effectively enhanced prospective memory performance in children with ADHD by specifically improving perceptual sensitivity (d’), with this beneficial effect maintained across different cognitive load levels without compromising ongoing task performance.

These findings provide novel insights into the cognitive mechanisms underlying prospective memory deficits in ADHD and offer evidence-based guidance for developing targeted intervention strategies that consider both internal cognitive resources and external environmental demands.

### 5.1. Theoretical Contributions

First, by investigating prospective memory (PM) performance in children with ADHD, this research advances our understanding of their PM processing characteristics and enables systematic examination of the dynamic tripartite relationships among cognitive load, implementation intention encoding, and prospective memory.

Second, we report the counterintuitive finding that elevated attentional load enhances PM performance in ADHD—a discovery that challenges prevailing theoretical models. Through Memory and Perception Theory (MPT) analysis, this phenomenon not only elucidates paradoxical cognitive behaviors but also extends the theoretical framework of MPT

Third, we demonstrate the practical and clinical value of implementation intention interventions. Incorporating “if-then” encoding significantly improved PM accuracy in children with ADHD, with notably divergent enhancement patterns observed under high versus low cognitive load conditions.

### 5.2. Practical Implications

Comparative analysis reveals significant prospective memory (PM) impairments in children with ADHD relative to typically developing peers, with the counterintuitive finding of enhanced PM performance under high cognitive load conditions informing a novel dynamic training protocol. This approach strategically leverages high-load contexts through cognitive load titration, context-embedded task implementation, and neurofeedback-guided modulation. Furthermore, implementation intention encoding markedly improves PM performance via ecological implementation templates (e.g., “If [classroom bell rings] → Then [submit homework]”), operationalized through sensory-anchored cueing, behavioral micro-scripting, and systematic generalization gradients to enhance ecological validity.

### 5.3. Limitations and Future Directions

This study has several limitations that warrant consideration. First, the experimental design included only low and high cognitive load conditions, omitting a no-load control group. This decision was intentional to focus on ecologically relevant load transitions (simulating daily light and demanding tasks, e.g., single vs. dual-task learning). However, this omission limits our ability to assess the absolute effect of load manipulation (e.g., whether high load enhances PM relative to baseline or merely lessens impairment). Future studies should include a no-load condition to clarify the true direction of load effects..Second, several paradigm-related limitations warrant consideration. Our PM task did not incorporate a filler or delay task between the instruction phase and the formal test. While this design choice was made to maintain task flow and minimize working memory demands for children with ADHD, it may have reduced the demand on truly ‘delayed’ intention retrieval, which is a core component of many naturalistic PM situations. Similarly, the relatively high frequency of PM trials (10 per block) within a short task, while necessary to obtain sufficient trials for reliable SDT analysis, may have increased the perceived salience of PM cues compared to real-world settings where PM actions typically occur less frequently. Future studies could systematically examine whether the presence of a retention interval or varying PM trial frequency moderates the effects observed here. Third, our study did not conduct subtype-specific analyses of PM deficits; future research should delineate whether impairment patterns in children with ADHD manifest differentially in event-based versus time-based PM paradigms. Furthermore, the examination of PM components remained undifferentiated. Subsequent investigations should dissect how distinct cognitive mechanisms—particularly the prospective component (cue detection and intention initiation) versus the retrospective component (content retrieval and action execution)—are modulated by varying cognitive load levels.

## Data Availability

All relevant data are within the manuscript and its Supporting Information files. Due to ethical restrictions related to participant confidentiality (the study involves children with ADHD and typically developing children), the raw data cannot be made publicly available. Data are available upon reasonable request from the corresponding author (Yixiao Pan), subject to approval from the Ethics Committee of Wenzhou Seventh People's Hospital.

## AUTHOR CONTRIBUTIONS

Conceptualization, Y.P. and J.H.; methodology, Y.P. and Z.L.; software, J.H.; validation, X.W. and Y.D.; formal analysis, Y.P., J.H. and Z.Y.; investigation, J.H., Z.L., X.W., Z.Y., Y.D. and Y.P.; resources, Y.P.; data curation, J.H. and Y.D.; writing—original draft preparation, J.H. and Y.P.; writing—review and editing, Z.L., X.W., Z.Y., Y.D. and Y.P.; visualization, J.H. and Y.P.; supervision, Y.P.; project administration, Y.P.; funding acquisition, Y.P. All authors have read and agreed to the published version of the manuscript.

## ACKNOWLEDGMENTS

Not applicable.

## DATA AVAILABILITY STATEMENT

The data that support the findings of this study are available from the corresponding author (Yixiao Pan) upon reasonable request. Data are not publicly available due to privacy restrictions of human participant data.

## ETHICS STATEMENT

The authors assert that all procedures contributing to this work comply with the ethical standards of the relevant national and institutional committees on human experimentation and with the Helsinki Declaration of 1975, as revised in 2008. All procedures involving human subjects/patients were approved by IRB in Wenzhou Seventh People’s Hospital (EC-KY-202408).

